# A mechanistic modeling approach to assessing the sensitivity of outcomes of water, sanitation, and hygiene interventions to local contexts and intervention factors

**DOI:** 10.1101/2024.03.09.24304020

**Authors:** Andrew F. Brouwer, Alicia N.M. Kraay, Mondal H. Zahid, Marisa C. Eisenberg, Matthew C. Freeman, Joseph N.S. Eisenberg

## Abstract

**Background:** Diarrheal disease is a leading cause of morbidity and mortality in young children. Water, sanitation, and hygiene (WASH) improvements have historically been responsible for major public health gains by reducing exposure to enteropathogens, but many individual interventions have failed to consistently reduce diarrheal disease burden. Analytical tools that can estimate the potential impacts of individual WASH improvements in specific contexts would support program managers and policymakers to set targets that would yield health gains.

**Methods:** To understand the impact of WASH improvements on diarrhea, we developed a disease transmission model to simulate an intervention trial with a single intervention. We accounted for contextual factors, including preexisting WASH conditions and baseline disease prevalence, as well as intervention WASH factors, including community coverage, compliance, efficacy, and the intervenable fraction of transmission. We illustrated the sensitivity of intervention effectiveness to the contextual and intervention factors in each of two scenarios in which a 50% reduction in disease was achieved through a different combination of factors (higher preexisting WASH conditions, compliance, and intervenable fraction vs higher intervention efficacy and community coverage).

**Results:** Achieving disease elimination depended on more than one factor, and factors that could be used to achieve disease elimination in one scenario could be ineffective in the other scenario. Community coverage interacted strongly with both the contextual and intervention factors. For example, the positive impact of increasing intervention community coverage increased non-linearly with increasing intervention compliance. Additionally, counterfactually improving the contextual preexisting WASH conditions could have a positive or negative effect on the intervention effectiveness, depending on the values of other factors.

**Conclusions:** When developing interventions, it is important to account for both contextual conditions and the intervention parameters. Our mechanistic modeling approach can provide guidance for developing locally specific policy recommendations.

## Introduction

Diarrheal disease is a leading cause of morbidity and mortality in young children, with an estimated 500,000 children under 5 years dying from diarrheal disease each year.^1–3^ Diarrheal disease is primarily caused by enteropathogens spread by fecal–oral pathways through contaminated environments, such as water, food, and fomites. Much of this burden is in low- and middle-income countries (LMICs) and among people living in poverty.^4^ Most enterpathogens are not good vaccine candidates, and those that are (e.g., rotavirus) can be hard to administer in the field (e.g., because of cold-chain requirements^5^) or suffer from differential effectiveness.^6^ Thus, preventive approaches for reducing enteric infections interventions are essential.

Historically, large-scale WASH improvements have been responsible for major public health gains by greatly reducing exposure to fecal pathogens, demonstrating the potential effectiveness of WASH in reducing the burden.^7,8^ Yet many trialed interventions, especially in the most disadvantaged areas where enteric infections are endemic, have failed to consistently reduce the burden of diarrhea disease.^9–15^ A recent meta-analysis of WASH intervention randomized controlled trials (RCTs) demonstrated that while WASH interventions can reduce diarrhea in children in low-resource settings overall,^16^ the heterogeneity across the aggregated trials is substantial, with many of the more recent, large-scale trials finding modest-to-null results.^9–15^

Difficulties in achieving consistent reduction of diarrheal burden are caused by multiple factors. First, local contexts can vary widely in terms of preexisting WASH conditions (i.e., WASH infrastructure in place prior to the intervention) and disease prevalence, among other factors. These differences have made it difficult to apply the results from studies conducted in one location to other locations. Second, interventions are imperfect. For example, 1) they may not block transmission along all transmission pathways (e.g., a water chlorination intervention will not reduce disease from exposure to animal feces or contaminated food), 2) the intervention coverage within the target population may not be sufficient to confer indirect protection, 3) the intervention may provide access to improved WASH but not ensure compliance, or 4) the direct efficacy of the provided interventions on reducing transmission to the users may be limited.^17,18^ Other factors are important as well, such as bias and inconsistency in reporting diarrhea and differences in the pathogens and taxa responsible for diarrheal disease in different locations.^19^

Analytical tools that can dynamically estimate the potential impacts of individual WASH improvements would support program managers and policymakers to set targets for investments to yield anticipated health gains. For example, with a given budget, should a program aim for greater coverage of an intervention or higher compliance, if the goal is health impact? Mechanistic transmission models can be enhanced to help implementors design optimal intervention strategies by accounting for location-specific contextual factors. One important strength of mechanistic approaches is their ability to generalize from available context-specific epidemiological findings to other contexts and counterfactual scenarios, and there is a need for tools that can generalize WASH trial results to other contexts.

Our objective was to develop a model to dynamically simulate diarrheal disease outcomes under various contextual and WASH intervention factors to understand which had the greatest impact on resulting disease burden. We previously developed a mechanistic model to simulate WASH trials and applied it to the WASH Benefits Bangladesh trial.^20^ Here, we aim to 1) demonstrate and estimate interactions between each of the contextual and intervention WASH factors and their impact on intervention efficacy and 2) increase the accessibility of the modeling framework for trialists and policymakers. This work will build our understanding of WASH interventions and improve the design of future trials.

## Methods

### WASH factors

In this analysis, we explore how effectiveness of a single intervention depends on six contextual or intervention WASH factors.

- *Preexisting WASH conditions*. We account for the fraction of the population that already has water, sanitation, and hygiene infrastructure comparable to that provided by the intervention.
- *Disease transmission potential*. We summarize disease transmission potential using the basic reproduction number *R*_0_. Note that because the baseline disease prevalence is determined by *R*_0_ (given the values of other the other factors), we will not independently vary the baseline disease prevalence in this analysis.
- *Intervention compliance*. We account for the fraction of participants assigned to an intervention that are actually using it. Compliance includes both fidelity (whether the intervention was delivered) and adherence (whether participants used the intervention).
- *Intervenable fraction of transmission*. Diarrheal disease pathogens are transmitted along multiple pathways, often summarized by an “F-diagram”: fluids, food, flies, fields, fauna, etc. Any individual intervention typically targets one or a few of these pathways, but not all of them, and each pathway is responsible for a different fraction of the total disease transmission potential. We account for how much transmission the intervention could prevent if it were perfectly efficacious and fully adopted. For trials that combine multiple interventions, the intervenable fraction can be thought of as the fraction of transmission that a combination of interventions could block.
- *Intervention efficacy*. Interventions do not perfectly prevent transmission along the pathways that they impact. We account for how much transmission (or shedding into the environment) the intervention prevents.
- *Community coverage*. In many trials, not everyone in the community is provided the interventions. We account for the fraction of the population that is enrolled in the trial.

Each of these factors is specifically accounted for in our transmission model, described below.

### Model

Our compartmental transmission model, denoted SISE-RCT, is a susceptible-infectious-susceptible (SIS) model with transmission through environmental (E) compartments. To approximate the outcomes of a RCT, we solve for the model’s steady state in an endemic setting.^20^ The SISE-RCT model accounts for the six mechanistic WASH factors outlined above that underlie WASH RCT results. In the case of a single intervention, the population is partitioned into individuals with regular exposure (those not enrolled or included in the intervention and those not compliant), and those with exposure or shedding attenuated by the intervention (those compliant with the intervention or an equivalent preexisting WASH condition). Susceptible and infectious individuals with regular exposure are designated *S*_-_ and *I*_-_, and those with exposure or shedding attenuated by the intervention are designated s_+_ and 1_+_. The intervention and control arms are simulated separately, and both the regular and attenuated exposure populations are modeled in both simulations, accounting for the fraction of population not enrolled in the study (ω), the fraction of the population with preexisting WASH conditions (*ρ*_0_), and intervention compliance (*ρ*). Individuals with regular exposure are either in the study but not compliant to the intervention (*ω*(1 – *ρ*)) or are not in the study and do not have preexisting WASH conditions ((1 – *ω*)(1 – *ρ*_0_)). Individuals with attenuated exposure are either in the study and compliant to the intervention (*ωρ*) or are not in the study but have preexisting WASH conditions ((1 – *ω*)*ρ*_0_). Hence, the population fractions of the attenuated and regular exposure populations are given by

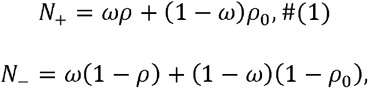

respectively.

Once infected, individuals clear the infection at rate *γ*. An environmental compartment is characterized by the shedding into the environment (*α*), the decay of pathogens in the environment (*ξ*), and the transmission of pathogens from the environment to susceptible individuals (*β*). For the single-intervention model, the environment is partitioned into the environmental pathway that is affected by the intervention *E*_1_, either in terms of shedding into or transmission from the environment, and the environmental pathway that is not affected by the intervention *E*_2_, with the same subscripts on *α*, (, and *β*. For example, *E*_1_ could be pathogens in water for an intervention that targets water, with *E*_2_ representing all other potential transmission pathways (e.g., fomites, food, etc). The relative magnitude of shedding into *E*_1_ and relative transmission from *E*_1_ for the attenuated compared to the exposed populations are given by 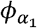and 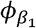, respectively.

The SISE-RCT parameters are given in Table 1, and a model diagram is given in Figure 1. The full equations are given below (Eqs 2). The two transmission terms *β*_1_*E*_1_ and *β*_2_*E*_2_ denote transmission from the environmental pathway attenuated by the intervention (*E*_1_) and from the environmental pathway not attenuated by the intervention (*E*_2_), respectively. The transmission term *β*_1_*E*_1_ is attenuated by 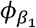 only for people in the attenuated exposure group (s_+_), and contamination of that environmental pathway is attenuated by 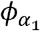 only for infectious people of that same group (1_+_). There is no attenuation of transmission to or shedding from the environmental pathway not affected by the intervention (*E*_2_). Parameters *ω, ρ*, and *ρ*_0_ do not show up in these equations but are accounted for in the constraints, as discussed below. For brevity, we omit the 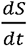 equations, each of which is given by 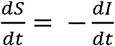 for the corresponding subpopulation.

**Table 1:** Parameters of the SISE-RCT model in two scenarios. The SISE-RCT model is a compartmental susceptible-infectious-susceptible (SIS) model with transmission through environmental (E) compartments and simulated to steady state to approximate an RCT. The intervention effectiveness in both scenarios in 50%, but the WASH parameters and baseline disease prevalence differ across scenarios.

| Parameter | Definition | Sensitivity range | Scenario 1 |  | Scenario 2 |  |
| --- | --- | --- | --- | --- | --- | --- |
|  |  |  | Baseline value | Disease elimination value | Baseline value | Disease elimination value |
| $\rho_0$ | Preexisting WASH conditions (fraction of individuals in the community with intervention-level WASH infrastructure) | 0–1 | 0.25 | 0.31 | 0 | — |
| $\rho$ | Compliance (fraction of individuals in intervention arm using intervention) | 0–1 | 0.75 | — | 0.50 | 0.92 |
| $R_0 = R_{0,1} + R_{0,2}$ | Transmission potential (basic reproduction number) | 1–2 | 1.25 | 1.20 | 1.25 | 1.12 |
| $\pi_c^*$ | Baseline disease prevalence | † | 6.4% | 2.8% | 20.0% | 10.7% |
| $R_{0,1}/(R_{0,1} + R_{0,2})$ | Intervenable fraction (fraction of transmission that the intervention could theoretically prevent) | 0–1 | 0.75 | 0.88 | 0.35 | 0.65 |
| $1 - \varphi_\alpha$ | Intervention efficacy for reducing shedding | — | 0 | — | 0 | — |
| $1 - \varphi_\beta$ | Intervention efficacy for reducing transmission | 0–1 | 0.75 | 0.98 | 0.83 | — |
| $\omega$ | Community coverage fraction (fraction of community included in the intervention trial) | 0–1 | 0.11 | 0.22 | 0.75 | — |

†: Baseline disease prevalence is a function of the transmission potential given the values of the other factors and was not varied independently of $R_0$ .

**Figure 1:**
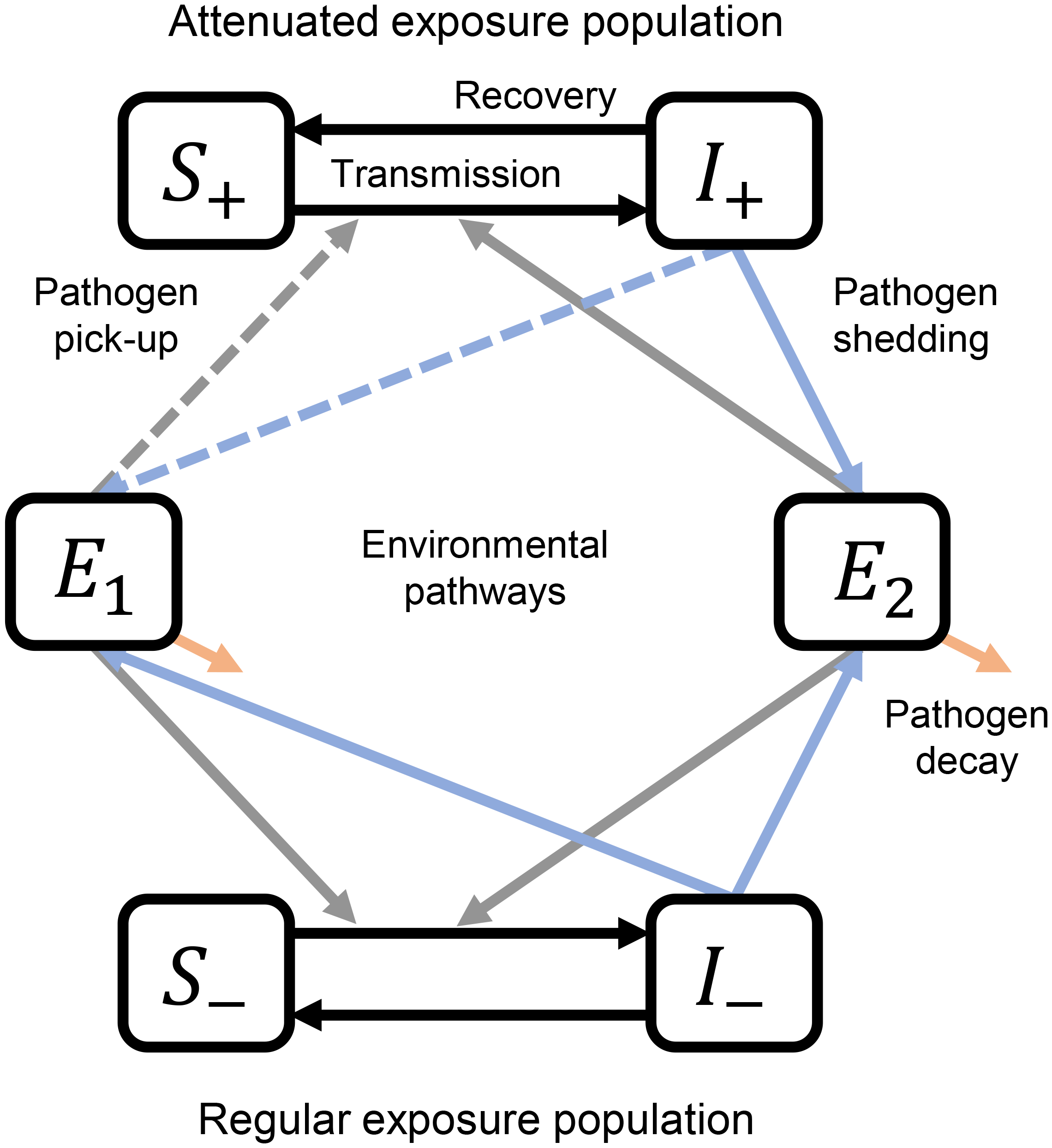
Single-intervention SISE-RCT model diagram with an attenuated exposure population and a regular exposure population interacting through shared environments. The SISE-RCT model is a compartmental susceptible-infectious-susceptible (SIS) model with transmission through environmental (E) compartments and simulated to steady state to approximate an RCT. The black lines denote infection and recovery, the blue lines denote shedding from infectious individuals into environmental compartments, the grey lines denote pick-up of pathogens from the environment by susceptible individuals, and the orange lines denote environmental pathogen decay. *S*_+_ and *I*_+_ denote susceptible and infectious fraction of the attenuate exposure population, and *S*_-_ and *I*_-_ denote susceptible and infectious fraction of the regular exposure population.

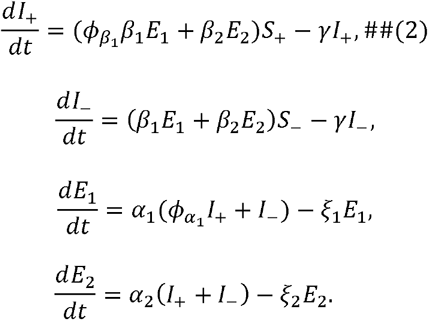

To find the steady state values (denoted by *) for the human compartments in the intervention arm, we set the above equations equal to 0 and simplify out the environmental compartments,

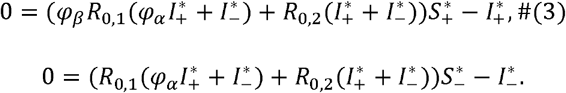

Here, 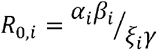 is the pathway-specific reproduction number for transmission through environment *E*_i_. For this specific model, the overall basic reproduction number is *R*_0_ = *R*_0,1_ + *R*_0,2_, denoting the sum of the transmission potential through the pathway affected by the intervention (*R*_0,1_) and the pathway not affected by the intervention (*R*_0,2_). The intervenable fraction (based on the strength of the transmission pathway targeted by the specific intervention) is *R*_0,1_/*R*_0_.

To get the steady states solutions for our four state variables 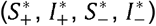, we solve the nonlinear system of equations (Eqs (3)) subject to the constraints 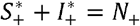 and 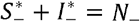, where *N*_+_ and *N*_-_ are given in Eqs (1). We solved this system using the `nleqslv` package in R. This approach is more computationally efficient than the differential equation simulation approach we used previously.^20^ We solve for the steady state in the control arm with the same parameters as the intervention arm except that *ρ* = *ρ*_0_.

The prevalence of disease in the population is denoted 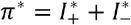 . The prevalence in the intervention arm π^*^ is compared to the prevalence 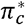 in the control arm. Then, intervention effectiveness (the RCT outcome) is defined as 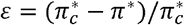, namely the fractional reduction in prevalence in the intervention arm relative to the control arm.

We investigated the sensitivity of the intervention effectiveness to each WASH factor. We first solved for the steady state solution for each of two scenarios with different sets of parameters, as listed in Table 1. Scenario 1 is characterized by a greater fraction of preexisting WASH conditions, compliance, and intervenable fraction, while Scenario 2 is characterized by a greater intervention efficacy and community coverage. The specific parameters in both scenarios were chosen to have 50% intervention effectiveness E but largely different values of the WASH contextual and intervention factors. The transmission potential was the same in both scenarios but the resulting baseline disease prevalence in Scenario 1 (6.4%) was much lower than that of Scenario 2 (20.9%) because of the differences in the other factors (particularly the preexisting conditions). The scenarios were chosen to demonstrate how sensitivity to the WASH factors might be different in different, plausible scenarios and are not intended to be representative of any specific intervention trial.

We varied each factor one at a time across the range of values given in Table 1, calculating the value needed to achieve disease elimination in that scenario. We also varied each pair of factors together (e.g., varying coverage and compliance together) to investigate potential interactions between factors. Only simulations with *ρ* > *ρ*_0_ and 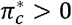 were included to avoid simulation of situations where the intervention reduced use of WASH or in which an intervention was applied to a system with no disease. This model has been made publicly available as a web app at https://umich-biostatistics.shinyapps.io/sise_rct/ and is included as supplementary material.

Note that the contextual factors, i.e., the preexisting WASH conditions and the transmission potential, are not modifiable in a real-world setting. In this analysis, changing these parameters represents the changing the location of the hypothetical trial and can help to reveal how the finds of a trial might generalize to other locations. While the sensitivity of intervention effectiveness to these parameters may be less relevant for trial planning in a specific location, it is important for developing a better understanding of the heterogeneity between trials and may also help to identify contexts where certain intervention approaches may be more effective than others.

## Results

The intervention effectiveness outcome E in Scenario 1, given by the parameters in Table 1, was 50%, with a steady-state prevalence of 6.4% in the control arm and 3.2% in the intervention arm. Disease elimination would have been achieved in this hypothetical intervention if 1) we increased the preexisting conditions so that 31% rather than 25% of the population already had comparable WASH infrastructure; 2) we reduced the disease potential transmission potential from *R*_0_=1.25 to *R*_0_=1.20, which is equivalent to reducing the baseline disease prevalence from 6.4% to 2.8%; 3) we increased the percentage of the total transmission that was blocked by the intervention from 75% to 88% (since there is an unknown, “true” value of the intervenable fraction, it may be more intuitive to think of this change as adding interventions until they target pathways responsible for 88% of the transmission potential); 4) we increased the efficacy of the intervention at reducing transmission from 75% to 88%; or 5) we increased the community coverage from 11% to 22%. Disease could not be eliminated by increasing intervention compliance from 75%, even to 100%.

The intervention effectiveness outcome E in Scenario 2, given by the parameters in Table 1, was also 50%, with a steady-state prevalence of 20.0% in the control arm and 10.0% in the intervention arm. Disease elimination would have been achieved in this hypothetical intervention if 1) we increased intervention compliance from 50% to 92%; 2) we reduced the disease potential transmission potential from *R*_0_=1.25 to *R*_0_=1.12, which is equivalent to reducing the baseline disease prevalence from 20.0% to 10.7%; or 3) we increased the percentage of the total transmission that was blocked by the intervention from 35% to 65%. The disease could not be eliminated with higher preexisting WASH conditions, higher efficacy, or higher community coverage.

The intervention effectiveness as a function of each pair of the six parameters is given in Figure 2 for Scenario 1 and in Figure 3 for Scenario 2, with each baseline scenario indicated by the white points. For many pairs of parameters, there was little evidence of an interaction between the factors (i.e., the contours of the heatmaps are approximately linear and parallel, except at extreme values). The primary exception to this pattern was coverage. In the inset in Figure 2, we show, as an illustration, the interaction between coverage and compliance on the intervention effectiveness. When coverage is low and compliance is high (point A), it is easier to increase intervention effectiveness by increasing coverage (gray arrow, moving along the x-axis), but when coverage is higher and compliance is low (point B), then it is easier to increase intervention effectiveness by increasing compliance (black arrow, moving along the y-axis). “Easier” here does not reflect cost or feasibility but only the result of a unit change for each individual parameter. Cost-effectiveness is outside of the scope of this work but could be explored in future analysis. Similarly, the coverage needed to achieve disease elimination depended non-linearly on each of the other factors.

**Figure 2:**
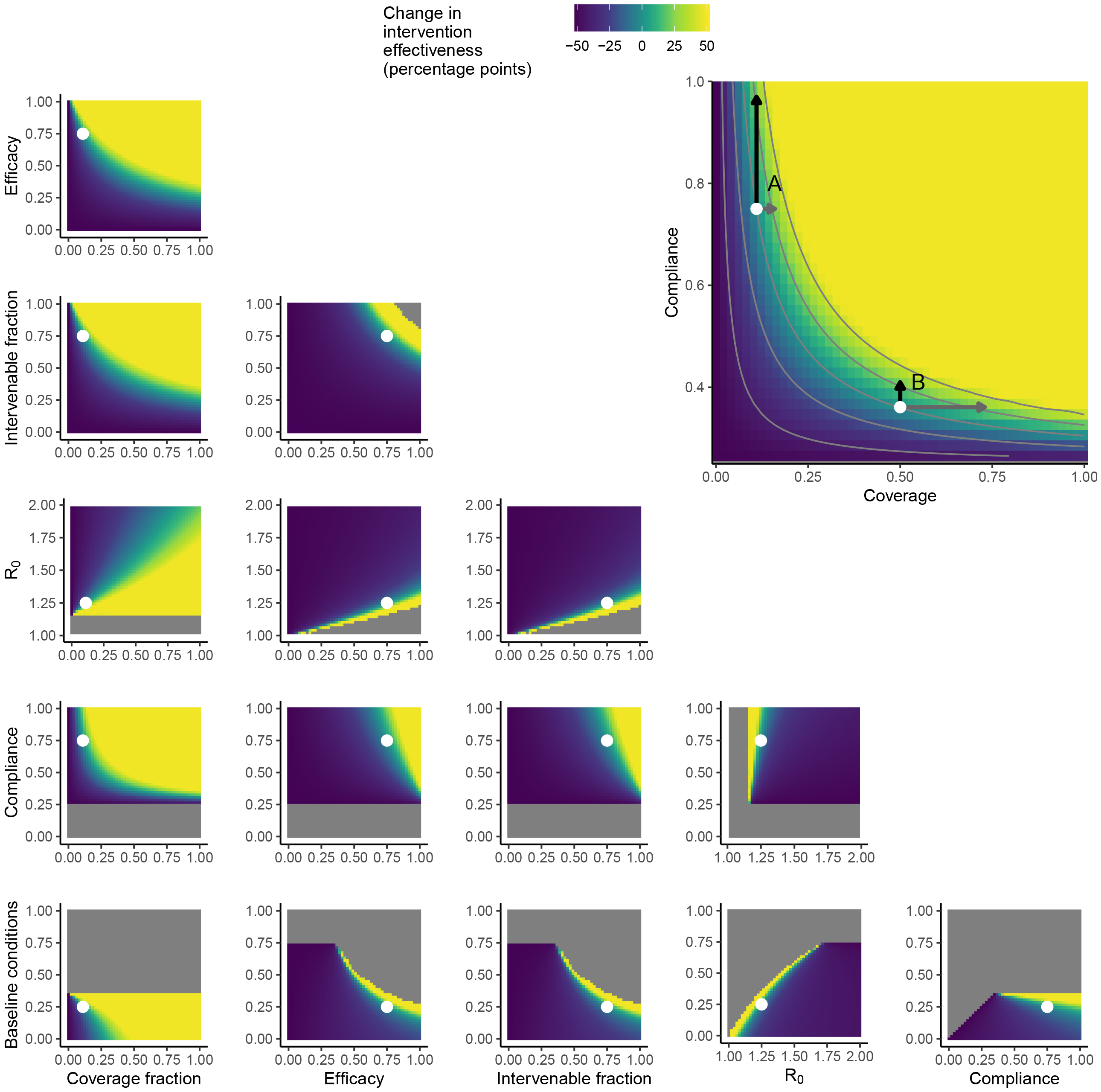
Intervention effectiveness as a function of WASH intervention factors in Scenario 1. The SISE-RCT model is a compartmental susceptible-infectious-susceptible (SIS) model with transmission through environmental (E) compartments and simulated to steady state to approximate an RCT. A single-intervention implementation of the model was simulated at the Scenario 1 baseline values given in Table 1 (indicated by the white points), and the heatmaps denote how intervention effectiveness depends on each pair of WASH factors. The six WASH factors are preexisting WASH conditions (fraction of individuals not enrolled in the intervention arm that are using preexisting infrastructure comparable to the intervention), compliance (fraction of individuals enrolled in the intervention arm that are using the intervention), disease transmission potential (summarize by the basic reproduction number *R*_0_), intervenable fraction of transmission (how much of the transmission could be prevented in a perfect intervention), intervention efficacy (fraction reduction in transmission or shedding when using the intervention), and the community coverage fraction (fraction of the population enrolled in the trial). The inset enlarges the compliance vs coverage plot and overlays contour lines to show the interaction between the two factors on intervention effectiveness. When coverage is low and compliance is high, it is easier to increase intervention effectiveness by increasing coverage, but when coverage is higher and compliance is low, then it is easier to increase intervention effectiveness by increasing compliance. WASH = water, sanitation, & hygiene.

**Figure 3:**
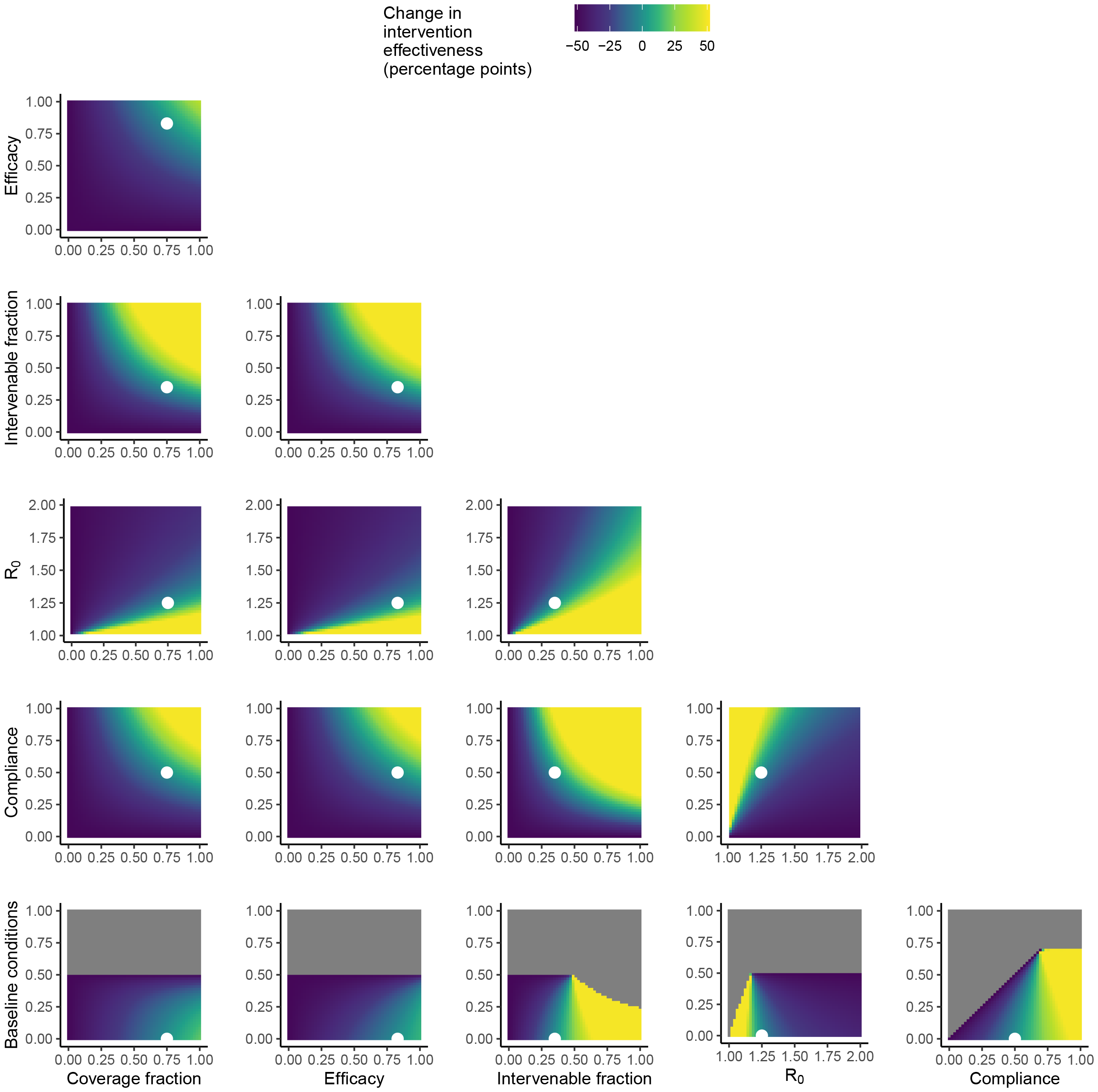
Intervention effectiveness as a function of WASH intervention factors in Scenario 2. Analogously to Figure 2, a single-intervention implementation of the model was simulated at the Scenario 2 baseline values given in Table 1 (and indicated by the white points), and the heatmaps denote how intervention effectiveness depends on each pair of WASH factors.

Increasing the fraction of the population with preexisting WASH conditions improved intervention effectiveness in Scenario 1 (Figure 2) but decreased intervention effectiveness in Scenario 2 (Figure 3). Increasing the fraction of the population with preexisting WASH conditions decreased prevalence in both the control and intervention arms, regardless of the specific scenario, but the *relative* reduction depended on the other WASH factors. In Scenario 2, for example, if the intervenable fraction were above 0.5 or if intervention compliance were above 0.75, then increasing baseline WASH conditions would result in increased intervention effectiveness.

The reader can explore the sensitivity of the model for other values of the WASH factors on the web app available via https://umich-biostatistics.shinyapps.io/sise_rct/ or using the code included as supplementary material.

## Discussion

We examined how the effectiveness of hypothetical single-intervention WASH RCTs depended on both contextual factors (baseline disease prevalence and preexisting WASH conditions) and intervention factors (community coverage, compliance, efficacy, and the intervenable fraction of transmission). Perhaps not surprisingly, the impact of changing one of some of these parameters was often highly dependent on the others. The effect of increasing community coverage, in particular, had a strong interaction with the other factors. For example, increasing the community coverage fraction could quickly lead to disease elimination if intervention compliance and efficacy were high, but have little impact if either were low. Our work demonstrates that it is important to understand the local, contextual conditions when developing relative priorities for an intervention. Our mechanistic modeling approach could allow for a tailored approach to designing interventions and WASH programs based on local conditions. For example, in some contexts with low baseline disease prevalence (like Scenario 1), substantial impacts might be achievable even with relatively low coverage. In contrast, in some contexts with high baseline disease prevalence (like Scenario 2), high coverage and compliance may be necessary to achieve strong efficacy.

Our findings offer a potential explanation for the high heterogeneity in the results of WASH intervention studies^16^ as well as the less-than-expected effectiveness of recent, large WASH intervention trials.^9–15^ An intervention that is effective in one location may be less effective in another location because of differences in the preexisting WASH infrastructure (e.g., the new location has unimproved latrines rather than open defecation) or differences in the disease pressure and baseline prevalence.

Additionally, there may be substantial differences in the distribution of enteropathogens in locations, as demonstrated by the MAL-ED and GEMS studies.^19,21,22^ These pathogens may use different transmission pathways, and, as a result, the intervenable transmission fraction for the intervention may be different in different locations.^4^ For example, norovirus is one of the hardest pathogens to control, as it can exploit multiple transmission pathways. Norovirus was particularly important as a cause of diarrheal disease at the MAL-ED study site in Nepal but was not detected at the site in India.^19^ Thus, an intervention blocking only one pathway might be less likely to reduce overall disease prevalence at the Nepal site, compared with India site. Moreover, the intervenable fraction may vary temporally within a site, as the dominant diarrheal pathogens may vary seasonally in their incidence. Continuing with the norovirus example, single-intervention effectiveness might also vary throughout the year and be less pronounced during cooler and wetter seasons, when norovirus is typically more prevalent.^23^

The strength of this analysis lies in the mechanistic framework that allows us to connect diarrheal disease outcomes in a WASH intervention context to the specific, measurable WASH factors that characterize the location and the intervention. Because we were interested in providing a basic understanding of the drivers of successful interventions, we decided to use hypothetical WASH factor values that were plausible but not specific to an existing trial. We also note that our models assume a steady state value for compliance; in practice, however, intervention compliance may decline over time.^24^ We plan to expand this sensitivity analysis to a full, multiple-intervention model and apply it to analyze real trials.

In the wake of the less-effective-than-expected large WASH intervention trial, a consensus group of WASH researchers called for a “pause for reflection” to re-evaluation the existing body of evidence.^17^ A recent meta-analysis has suggested that WASH is effective at reducing diarrheal disease, though the outcomes are highly heterogeneous.^16^ Our mechanistic modeling framework is another approach that is well-suited to re-evaluating existing evidence and generating hypotheses for causal explanations of the results of these trials. Ultimately, our work will help to provide evidence for developing locally specific policy recommendations and programmatic targets and for designing the next-generation WASH interventions.^18,25,26^

## Contributors

JNSE, MCE, MCF, and AFB conceived of the study. JNSE and MCF secured funding for the study. AFB, MCE, and JNSE developed the model. AFB wrote and implemented the software code, completed formal analysis and visualization, and curated the data and code. AFB wrote the original draft with input from JNSE, MCF, and ANMK. All authors reviewed and edited the manuscript. All authors had full access to all study data.

## Acknowledgements

This work was funded by the Bill & Melinda Gates Foundation (grant INV-005081) and the National Science Foundation (grant DMS-1853032). Study sponsors had no role in the study design, the analysis or interpretation of the results, the writing of the report, or in the decision to submit the paper for publication.

## Data availability statement

No data are associated with this article. The code is included as supplemental material. The SISE-RCT web app with the single-intervention model is available at https://umich-biostatistics.shinyapps.io/sise_rct/.

