## Supplementary material for "A mechanistic modeling approach to assessing the sensitivity of outcomes of water, sanitation, and hygiene interventions to local contexts and intervention factors": SISE-RCT.R

```
library(deSolve)
library(ggplot2)
library(ggpubr)
library(scales)
library(viridis)
library(nleqslv)


###################################################################################
#Model functions
model = function(x, model_par){
  
  x= abs(x)
  
  #Effectiveness
  pi_alpha = model_par[1]
  pi_beta = model_par[2]
  #Relative value
  phi_alpha = 1 - pi_alpha
  phi_beta = 1 - pi_beta
  
  #R0 N = intervenable, O= other
  R0_N = model_par[3]
  R0_O = model_par[4]
  
  par_baseline_condition = model_par[5]
  par_intervention_compliance = model_par[6]
  coverage=model_par[7]
  
  total_SI = (1-par_baseline_condition)*(1-coverage) + (1-par_intervention_compliance)*coverage
  total_SI_N = par_baseline_condition*(1-coverage) + par_intervention_compliance * coverage
  
  I = x[1]
  I_N = x[2]
  
  S = x[3]
  S_N = x[4]
  
  y = numeric(length(x))
  y[1]  =  S * ( R0_N * (I + I_N*phi_alpha) + R0_O * (I + I_N)) -  I
  y[2]  =  S_N * (phi_beta * R0_N * (I + I_N*phi_alpha) + 
                    R0_O * (I + I_N)) -  I_N
  
  y[3] =   I + S - total_SI
  y[4] =   I_N + S_N - total_SI_N
  
  return(y)
  
}

model_Jac = function(x, model_par){
  
  x=abs(x)
  
  #Effectiveness
  pi_alpha = model_par[1]
  pi_beta = model_par[2]
  #Relative value
  phi_alpha = 1 - pi_alpha
  phi_beta = 1 - pi_beta
  
  #R0 N = intervenable, O= other
  R0_N = model_par[3]
  R0_O = model_par[4]
  
  par_baseline_condition = model_par[5]
  par_intervention_compliance = model_par[6]
  coverage=par[7]
  
  I = x[1]
  I_N = x[2]
  
  S = x[3]
  S_N = x[4]
  
  Df = matrix(NA, length(x), length(x))
  Df[1,1]= S * (R0_N  + R0_O) -1
  Df[1,2]= S * (R0_N*phi_alpha  + R0_O)
  Df[1,3]= (R0_N * (I + I_N*phi_alpha) + R0_O * (I + I_N))
  Df[1,4]= 0
  
  Df[2,1]= S_N * (phi_beta * R0_N  + R0_O)
  Df[2,2]= S_N * (phi_beta * phi_alpha * R0_N  + R0_O) - 1
  Df[2,3]= 0
  Df[2,4]= (phi_beta * R0_N * (I + I_N*phi_alpha) + R0_O * (I + I_N))
  
  Df[3,1]= 1
  Df[3,2]= 0
  Df[3,3]= 1
  Df[3,4]= 0
  
  Df[4,1]= 0
  Df[4,2]= 1
  Df[4,3]= 0
  Df[4,4]= 1
  
  return(Df)
  
}

#Simulate prevalence as a function of the parameters
simulate = function(par){
  
  par_baseline_condition = par[1]
  par_intervention_compliance = par[2]
  R0 = par[3]
  R0_ratio = par[4]
  efficacy_alpha = par[5]
  efficacy_beta= par[6]
  coverage=par[7]
  
  rho_vec = c(1-par_intervention_compliance, par_intervention_compliance)
  baseline_adherence_vec = c(1-par_baseline_condition, par_baseline_condition)
  
  total_SI = (1-par_baseline_condition)*(1-coverage) + (1-par_intervention_compliance)*coverage
  total_SI_N = par_baseline_condition*(1-coverage) + par_intervention_compliance * coverage
  
  prev= 1-1/R0 #guess
  # x0 =c(I, I_N, S, S_N)
  x0 = c(prev*total_SI,prev*total_SI_N,(1-prev)*total_SI,(1-prev)*total_SI_N)
  
  R0_N= R0*R0_ratio
  R0_O = R0*(1-R0_ratio)
  model_par = c(efficacy_alpha,efficacy_beta, R0_N,R0_O,
                par_baseline_condition,par_intervention_compliance,
                coverage)
  
  steady_state = abs(nleqslv(x=x0,fn=model,jac = model_Jac, method="Broyden",model_par=model_par)$x)
  
  prevalence  = sum(steady_state[1:2])
  
  return(prevalence)
}


###############################
#Intervention simulation
###############################

#Baseline conditions. What fraction of population already has WASH infrastructure similar to the intervention?
#Accept values between 0 and 100
par_baseline_condition = 25#% 

#Intervention compliance. What fraction of the intervention group receives and uses the already has WASH infrastructure similar to the intervention?
#Accept values between 0 and 100
par_intervention_compliance = 75#%

#Basic reproduction number. Controls the burden of disease.
#Accept values between 1 and 2
R0=1.25

#Intervenable fraction. How much of the transmission could be controlled if the intervention was perfectly effective?
#Accept values between 0 and 1
R0_ratio = 0.75

#Does the intervention reduce shedding into the environment or transmission from the environment to people?
#Toggle between "Shedding" (flag=0) and "Transmission" (flag=1)
flag = 1

#Intervention efficacy. What percent of shedding/transmission is prevented by the intervention?
#Accept values between 0 and 100
efficacy = 75#%

#Efficacy at reducing infection. What percent of potential transmission is prevented by the intervention?
#Accept values between 0 and 100
efficacy_beta= 75#%

#What percent of the population is included in the intervention?
#Accept values between 0 and 100
coverage = 10.75 #Display: %

#Set parameters based on the above
# par = c(par_baseline_condition/100, par_intervention_compliance/100,
#         R0, R0_ratio, (1-flag)*efficacy/100,  flag*efficacy/100, coverage/100)

#Or choose one of these scenarios
#Scenario 1
par=c(0.25, 0.75, 1.25, 0.75, 0, 0.75, 0.1075)

#Scenario 2
# par = c(0, 0.50, 1.25, 0.35, 0, 0.83, 0.75)

#Baseline control
prev_control = simulate(c(par[1:6],0))
#Baseline intervention
prev_intervention = simulate(par)
#Baseline max intervention
prev_intervention_max = simulate(c(par[1],1,par[3:4],(1-flag),flag,par[7]))


data_plot1=as.data.frame(cbind(c("Control","Control","Intervention","Intervention"),
                               100*c(prev_intervention_max,prev_control-prev_intervention_max,
                                     prev_intervention_max,prev_intervention-prev_intervention_max),
                               c("Non-intervenable","Intervenable","Non-intervenable","Intervenable")))
colnames(data_plot1)=c("Arm","Prevalence","Transmission")
data_plot1$Prevalence = as.numeric(data_plot1$Prevalence )

relative_risk_actual = prev_intervention/prev_control
intervention_effectiveness_actual = 1- relative_risk_actual

#Compare prevalence in the control and intervention arms
ggplot(data_plot1,aes(x=Arm,y=Prevalence,fill=Transmission))+
  geom_col()+theme_classic()+ylab("Prevalence (%)")+
  scale_fill_manual(values=c("grey75","grey25"))

#Prevalence in control arm
round(prev_control*100,digits=1)
#Prevalence in intervention arm
round(prev_intervention*100,digits=1)
#Intervention effectiveness. What percent of disease is prevented by the intervention?
round(intervention_effectiveness_actual * 100,digits=1) #%

###############################
#Sensitivity analysis
###############################
#Color limits for display
col_limits = c(-52,52)

fill_matrix=function(par0,factor1,factor2,res){
  flag = par0[5]==0
  
  matrix = matrix(NA, res*res,3)
  
  for (i in 1:res){
    for (j in 1:res){
      par=par0
      
      if (factor1=="conditions"){par[1] = seq(0,1,length.out = res)[i]
      factor1_temp=par[1]}
      if (factor1=="compliance"){par[2] = seq(0,1,length.out = res)[i]
      factor1_temp=par[2]}
      if (factor1=="R0"){par[3] = seq(1,2,length.out = res)[i]
      factor1_temp=par[3]}
      if (factor1=="completeness"){par[4] = seq(0,1,length.out = res)[i]
      factor1_temp=par[4]}
      if (factor1=="efficacy"){par[5] = seq(0,1,length.out = res)[i]*(1-flag)
      par[6] = seq(0,1,length.out = res)[i]*flag
      factor1_temp=seq(0,1,length.out = res)[i]}
      if (factor1=="coverage"){par[7] = seq(0,1,length.out = res)[i]
      factor1_temp=par[7]}
      
      if (factor2=="conditions"){par[1] = seq(0,1,length.out = res)[j]
      factor2_temp=par[1]}
      if (factor2=="compliance"){par[2] = seq(0,1,length.out = res)[j]
      factor2_temp=par[2]}
      if (factor2=="R0"){par[3] = seq(1,2,length.out = res)[j]
      factor2_temp=par[3]}
      if (factor2=="completeness"){par[4] = seq(0,1,length.out = res)[j]
      factor2_temp=par[4]}
      if (factor2=="efficacy"){par[5] = seq(0,1,length.out = res)[j]*(1-flag)
      par[6] = seq(0,1,length.out = res)[j]*flag
      factor2_temp=seq(0,1,length.out = res)[j]}
      if (factor2=="coverage"){par[7] = seq(0,1,length.out = res)[j]
      factor2_temp=par[7]}
      
      if (par[1]>par[2]){
        change_intervention_effectiveness = NA
      }else{
        prev_control = simulate(c(par[1:6],0))
        if (prev_control<1E-4){
          change_intervention_effectiveness = NA
        }else{ #This got fixed
          prev_intervention = simulate(par)
          relative_risk_counterfactual = prev_intervention/prev_control
          intervention_effectiveness_counterfactual = 1- relative_risk_counterfactual
          
          change_intervention_effectiveness = 100*(intervention_effectiveness_counterfactual-intervention_effectiveness_actual)}
      }
      matrix[j+(i-1)*res,]=c(factor2_temp, factor1_temp, change_intervention_effectiveness)
    }
  }
  matrix = as.data.frame(matrix)
  matrix$xlab=factor2
  matrix$ylab=factor1
  colnames(matrix)=c("x","y","value","xlab","ylab")
  return(matrix)
}

#Resolution
res = 50

#conditions, compliance
matrix1= fill_matrix(par,"conditions","compliance",res)
p1= ggplot()+
  geom_raster(data=matrix1, aes(x=x,y=y,fill=value))+
  annotate("point",x=par[2],y=par[1],pch=19,col="white",size=3)+
  scale_fill_gradientn(colors = viridis(10),values=rescale(c(-1,-0.5,-0.25,0,0.25,0.5,1)),limits=col_limits)+
  guides(fill=guide_colorbar(title="Change in\nintervention\neffectiveness\n(percentage points)        "))+
  xlab("Compliance")+ylab("")+#ylab("Baseline conditions")+
  theme_classic()

#conditions, R0
matrix2= fill_matrix(par,"conditions","R0",res)
p2= ggplot()+
  geom_raster(data=matrix2, aes(x=x,y=y,fill=value))+
  annotate("point",x=par[3],y=par[1],pch=19,col="white",size=3)+
  scale_fill_gradientn(colors = viridis(10),values=rescale(c(-1,-0.5,-0.25,0,0.25,0.5,1)),limits=col_limits)+
  guides(fill=guide_colorbar(title="Change in\nintervention\neffectiveness\n(percentage points)        "))+
  xlab(expression(R[0]))+ylab("")+#+ylab("Baseline conditions")+
  theme_classic()

#conditions, completeness
matrix3= fill_matrix(par,"conditions","completeness",res)
p3= ggplot()+
  geom_raster(data=matrix3, aes(x=x,y=y,fill=value))+
  annotate("point",x=par[4],y=par[1],pch=19,col="white",size=3)+
  scale_fill_gradientn(colors = viridis(10),values=rescale(c(-1,-0.5,-0.25,0,0.25,0.5,1)),limits=col_limits)+
  guides(fill=guide_colorbar(title="Change in\nintervention\neffectiveness\n(percentage points)        "))+
  xlab("Intervenable fraction")+ylab("")+#+ylab("Baseline conditions")+
  theme_classic()

#conditions, efficacy
matrix4= fill_matrix(par,"conditions","efficacy",res)
p4= ggplot()+
  geom_raster(data=matrix4, aes(x=x,y=y,fill=value))+
  annotate("point",x=c(par[5],par[6])[flag+1],y=par[1],pch=19,col="white",size=3)+
  scale_fill_gradientn(colors = viridis(10),values=rescale(c(-1,-0.5,-0.25,0,0.25,0.5,1)),limits=col_limits)+
  guides(fill=guide_colorbar(title="Change in\nintervention\neffectiveness\n(percentage points)        "))+
  xlab("Efficacy")+ylab("")+#+ylab("Baseline conditions")+
  theme_classic()

#conditions, coverage
matrix5= fill_matrix(par,"conditions","coverage",res)
p5= ggplot()+
  geom_raster(data=matrix5, aes(x=x,y=y,fill=value))+
  annotate("point",x=par[7],y=par[1],pch=19,col="white",size=3)+
  scale_fill_gradientn(colors = viridis(10),values=rescale(c(-1,-0.5,-0.25,0,0.25,0.5,1)),limits=col_limits)+
  guides(fill=guide_colorbar(title="Change in\nintervention\neffectiveness\n(percentage points)        "))+
  xlab("Coverage fraction")+ylab("Baseline conditions")+
  theme_classic()

#compliance, R0
matrix6= fill_matrix(par,"compliance","R0",res)
p6= ggplot()+
  geom_raster(data=matrix6, aes(x=x,y=y,fill=value))+
  annotate("point",x=par[3],y=par[2],pch=19,col="white",size=3)+
  scale_fill_gradientn(colors = viridis(10),values=rescale(c(-1,-0.5,-0.25,0,0.25,0.5,1)),limits=col_limits)+
  guides(fill=guide_colorbar(title="Change in\nintervention\neffectiveness\n(percentage points)        "))+
  # xlab(expression(R[0]))+ylab("Compliance")+
  xlab("")+ylab("")+
  scale_x_continuous(limits=c(1,2),breaks=c(1,1.25,1.5,1.75,2))+
  theme_classic()

#compliance, completeness
matrix7= fill_matrix(par,"compliance","completeness",res)
p7= ggplot()+
  geom_raster(data=matrix7, aes(x=x,y=y,fill=value))+
  annotate("point",x=par[4],y=par[2],pch=19,col="white",size=3)+
  scale_fill_gradientn(colors = viridis(10),values=rescale(c(-1,-0.5,-0.25,0,0.25,0.5,1)),limits=col_limits)+
  guides(fill=guide_colorbar(title="Change in\nintervention\neffectiveness\n(percentage points)        "))+
  # xlab("Intervenable fraction")+ylab("Compliance")+
  xlab("")+ylab("")+
  theme_classic()

#compliance, efficacy
matrix8= fill_matrix(par,"compliance","efficacy",res)
p8= ggplot()+
  geom_raster(data=matrix8, aes(x=x,y=y,fill=value))+
  annotate("point",x=c(par[5],par[6])[flag+1],y=par[2],pch=19,col="white",size=3)+
  scale_fill_gradientn(colors = viridis(10),values=rescale(c(-1,-0.5,-0.25,0,0.25,0.5,1)),limits=col_limits)+
  guides(fill=guide_colorbar(title="Change in\nintervention\neffectiveness\n(percentage points)        "))+
  # xlab("Efficacy")+ylab("Compliance")+
  xlab("")+ylab("")+
  theme_classic()

#compliance, coverage
matrix9= fill_matrix(par,"compliance","coverage",res)
p9= ggplot()+
  geom_raster(data=matrix9, aes(x=x,y=y,fill=value))+
  annotate("point",x=par[7],y=par[2],pch=19,col="white",size=3)+
  scale_fill_gradientn(colors = viridis(10),values=rescale(c(-1,-0.5,-0.25,0,0.25,0.5,1)),limits=col_limits)+
  guides(fill=guide_colorbar(title="Change in\nintervention\neffectiveness\n(percentage points)        "))+
  # xlab("Coverage fraction")
  xlab("")+ylab("Compliance")+
  theme_classic()

#R0, completeness
matrix10= fill_matrix(par,"R0","completeness",res)
p10= ggplot()+
  geom_raster(data=matrix10, aes(x=x,y=y,fill=value))+
  annotate("point",x=par[4],y=par[3],pch=19,col="white",size=3)+
  scale_fill_gradientn(colors = viridis(10),values=rescale(c(-1,-0.5,-0.25,0,0.25,0.5,1)),limits=col_limits)+
  guides(fill=guide_colorbar(title="Change in\nintervention\neffectiveness\n(percentage points)        "))+
  # xlab("Intervenable fraction")+ylab(expression(R[0]))+
  scale_y_continuous(limits=c(1,2),breaks=c(1,1.25,1.5,1.75,2))+
  xlab("")+ylab("")+
  theme_classic()

#R0, efficacy
matrix11= fill_matrix(par,"R0","efficacy",res)
p11= ggplot()+
  geom_raster(data=matrix11, aes(x=x,y=y,fill=value))+
  annotate("point",x=c(par[5],par[6])[flag+1],y=par[3],pch=19,col="white",size=3)+
  scale_fill_gradientn(colors = viridis(10),values=rescale(c(-1,-0.5,-0.25,0,0.25,0.5,1)),limits=col_limits)+
  guides(fill=guide_colorbar(title="Change in\nintervention\neffectiveness\n(percentage points)        "))+
  # xlab("Efficacy")+ylab(expression(R[0]))+
  scale_y_continuous(limits=c(1,2),breaks=c(1,1.25,1.5,1.75,2))+
  xlab("")+ylab("")+
  theme_classic()

#R0, coverage
matrix12= fill_matrix(par,"R0","coverage",res)
p12= ggplot()+
  geom_raster(data=matrix12, aes(x=x,y=y,fill=value))+
  annotate("point",x=par[7],y=par[3],pch=19,col="white",size=3)+
  scale_fill_gradientn(colors = viridis(10),values=rescale(c(-1,-0.5,-0.25,0,0.25,0.5,1)),limits=col_limits)+
  guides(fill=guide_colorbar(title="Change in\nintervention\neffectiveness\n(percentage points)        "))+
  # xlab("Coverage fraction")
  xlab("")+ylab(expression(R[0]))+
  scale_y_continuous(limits=c(1,2),breaks=c(1,1.25,1.5,1.75,2))+
  theme_classic()

#completeness, efficacy
matrix13= fill_matrix(par,"completeness","efficacy",res)
p13= ggplot()+
  geom_raster(data=matrix13, aes(x=x,y=y,fill=value))+
  annotate("point",x=c(par[5],par[6])[flag+1],y=par[4],pch=19,col="white",size=3)+
  scale_fill_gradientn(colors = viridis(10),values=rescale(c(-1,-0.5,-0.25,0,0.25,0.5,1)),limits=col_limits)+
  guides(fill=guide_colorbar(title="Change in\nintervention\neffectiveness\n(percentage points)        "))+
  # xlab("Efficacy")+ylab("Intervenable fraction")+
  xlab("")+ylab("")+
  theme_classic()

#completeness, coverage
matrix14= fill_matrix(par,"completeness","coverage",res)
p14= ggplot()+
  geom_raster(data=matrix14, aes(x=x,y=y,fill=value))+
  annotate("point",x=par[7],y=par[4],pch=19,col="white",size=3)+
  scale_fill_gradientn(colors = viridis(10),values=rescale(c(-1,-0.5,-0.25,0,0.25,0.5,1)),limits=col_limits)+
  guides(fill=guide_colorbar(title="Change in\nintervention\neffectiveness\n(percentage points)        "))+
  # xlab("Coverage fraction")
  xlab("")+ylab("Intervenable fraction")+
  theme_classic()

#efficacy, coverage
matrix15= fill_matrix(par,"efficacy","coverage",res)
p15= ggplot()+
  geom_raster(data=matrix15, aes(x=x,y=y,fill=value))+
  annotate("point",x=par[7],y=c(par[5],par[6])[flag+1],pch=19,col="white",size=3)+
  scale_fill_gradientn(colors = viridis(10),values=rescale(c(-1,-0.5,-0.25,0,0.25,0.5,1)),limits=col_limits)+
  guides(fill=guide_colorbar(title="Change in\nintervention\neffectiveness\n(percentage points)        "))+
  # xlab("Coverage fraction")
  xlab("")+ylab("Efficacy")+
  theme_classic()

#Arrange plots
p=ggarrange(p15,NULL,NULL,NULL,NULL,
            p14,p13,NULL,NULL,NULL,
            p12,p11,p10,NULL,NULL,
            p9,p8,p7,p6,NULL,
            p5,p4,p3,p2,p1,
            ncol=5,nrow=5,common.legend = TRUE,legend="top")
p=p+annotation_custom(
  ggplotGrob(p_inset),
  xmin = 0.6, xmax = 1, ymin = 0.53, ymax = 0.93)
p
```
